# Glucagon-Like Peptide-1 Receptor Agonists and Risk of Parkinsonian Disorders in Patients with Obesity

**DOI:** 10.64898/2025.11.24.25340763

**Authors:** Pareeta Kotecha, Yao An Lee, Michael F Presti, Yi Guo, Jiang Bian, Jingchuan Guo

## Abstract

**Introduction:** Parkinsonism encompasses neurodegenerative disorders characterized by rigidity, tremor, and bradykinesia, with Parkinson’s disease (PD) accounting for most cases. Emerging evidence suggests that metabolic dysfunction, including insulin resistance and neuroinflammation, may contribute to PD pathogenesis. Glucagon-like peptide-1 receptor agonists (GLP-1RAs), approved for type 2 diabetes (T2D) and obesity, demonstrate neuroprotective effects in preclinical PD models. However, real-world evidence in individuals without T2D is limited. This study evaluated whether GLP-1RA use among adults with obesity or overweight is associated with the risk of PD and Parkinsonian disorders.

**Methods:** We conducted a retrospective cohort study using 2014-2024 electronic health record data from the OneFlorida+ Network. Adults with obesity (BMI ≥30 kg/m^2^), overweight (BMI 25-29.9 kg/m^2^) with a weight-related comorbidity, or an obesity diagnosis were eligible. Exclusions included age <50 years, <30 days of follow-up, baseline PD or Parkinsonism, anti-Parkinson medication use, or T2D. GLP-1RA users were matched 1:1 to non-users using time-conditional propensity scores. Outcomes were a composite of (1) PD/Parkinsonism and (2) any Parkinsonian-related disorder, including PD, Parkinsonism, Lewy body dementias, and multiple system atrophy. Cox proportional hazards models estimated hazard ratios (HRs) and 95% confidence intervals (CIs).

**Results:** The matched cohort included 11,683 GLP-1RA users and 11,683 non-users with balanced characteristics and a median follow-up of 286 days. GLP-1RA use was not associated with reduced risk of PD or Parkinsonism (22 vs. 33 events; HR 0.70, 95% CI 0.41-1.22) or any Parkinsonian-related disorder (98 vs. 108 events; HR 0.96, 95% CI 0.73-1.27). Findings were consistent in subgroup and sensitivity analyses.

**Conclusion:** Among adults with obesity or overweight with a weight related condition, there was no difference in the risk of PD or Parkinsonian disorders between GLP-1 RA users and non-users. Longer follow-up and clinical trials are needed to clarify potential neuroprotective effects across populations.

## Introduction

Parkinsonism encompasses a group of neurodegenerative disorders characterized by rigidity, tremor, and bradykinesia, with Parkinson disease (PD) accounting for most cases.^1^ PD is the second most common neurodegenerative disease and affects nearly one million people in the United States.^2^ In addition to aging, genetics, and environmental exposures, metabolic dysfunction, particularly insulin resistance, mitochondrial impairment, and neuroinflammation has been implicated in PD pathogenesis.^3^ Glucagon-like peptide-1 receptor agonists (GLP-1RAs), approved for type 2 diabetes (T2D) and obesity treatment, show neuroprotective effects in preclinical PD models.^4^ However, real-world evidence has focused largely on patients with T2D. This study evaluates whether GLP-1RA use among adults with obesity or overweight and weight related condition and without T2D is associated with the risk of PD and parkinsonian disorders.

## Methods

We conducted a retrospective cohort study using 2014-2024 electronic health record (EHR) data from the OneFlorida+ Clinical Research Network, which includes more than 20 million patients across Florida, Georgia, and Alabama. Adults with recorded obesity (BMI ≥30 kg/m^2^), overweight with a weight-related comorbidity (BMI 25-29.9 kg/m^2^), or an ICD (9 and 10) diagnosis of obesity were eligible. Patients were required to initiate a GLP-1RA after their first obesity-related encounter. Individuals aged <50 years; those with <30 days of follow-up; or with baseline PD, parkinsonism, multiple system atrophy, Lewy body dementia, anti-Parkinson medication use, or T2D were excluded.

GLP-1RA users were matched 1:1 to non-users using time-dependent propensity scores based on the closest clinical encounter around GLP-1RA initiation. Outcomes included (i) composite outcome of PD/parkinsonism and (ii) Any: PD/ parkinsonism /Lewy body dementia/multiple system atrophy. The patients were followed until the first occurrence of any outcome within the composite outcome, death, last encounter in EHR or January 31, 2024, whichever occurred first. Subgroup and sensitivity analyses assessed robustness across demographic strata and minimum follow-up thresholds by including those with at least i) 6 months and ii)1 year of follow-up. Hazard ratios (HRs) and 95% CIs were estimated using Cox proportional hazards models. The study followed STROBE guidelines and was approved by the University of Florida IRB.

## Results

After time-dependent propensity score matching, 11,683 GLP-1RA users were matched to 11,683 non-users with well-balanced baseline characteristics (Table), with a median follow-up of 286 days (IQR: 154-490 days). GLP-1RA use was not associated with a lower risk of either parkinsonian disorder outcomes. For the composite outcome of PD and parkinsonism, 22 events (0.2%) occurred among GLP-1RA users and 33 events (0.3%) among nonusers (HR, 0.70 [95% CI, 0.41-1.22]). For any Parkinsonian related outcome, a total of 98 events (0.8%) occurred among users and 108 events (0.9%) among non-users (HR, 0.96 [95% CI, 0.73-1.27]). Subgroup analyses showed consistent findings across age, race and ethnicity, and drug class, although women demonstrated a lower risk for both outcomes with GLP-1RA use, whereas no protective association was observed among men. Sensitivity analyses requiring at least 6 months or 1 year of follow-up yielded similar results demonstrating no significant effect on the risk of Parkinson related disorders between GLP-1 RA users and non-users (Figure).

## Discussion

In this large cohort of adults with obesity or overweight with a weight related condition, GLP-1RA use was not associated with a reduced risk of parkinsonian disorders. The findings were consistent across two composite outcomes, subgroup and sensitivity analyses. Our findings were consistent with phase 3 trial data showing no disease-modifying effect of exenatide in patients with established PD.^5^ Similarly, a real-world study restricted to patients with obesity (and T2D) did not show statistically significant reductions in PD risk.^6^ A notable exception was the lower risk observed among women, which may be explained by sex differences in dopaminergic pathways, inflammatory responses, or GLP-1 receptor expression, but the findings should be interpreted with caution given the small number of events.^4^ This study has limitations, including short follow-up for disorders with long prodromal phases, potential residual confounding, and smaller subgroups. Long term studies and clinical trials are needed to clarify whether GLP-1 RAs influence risk of parkinsonian related disorders.

**Table:**
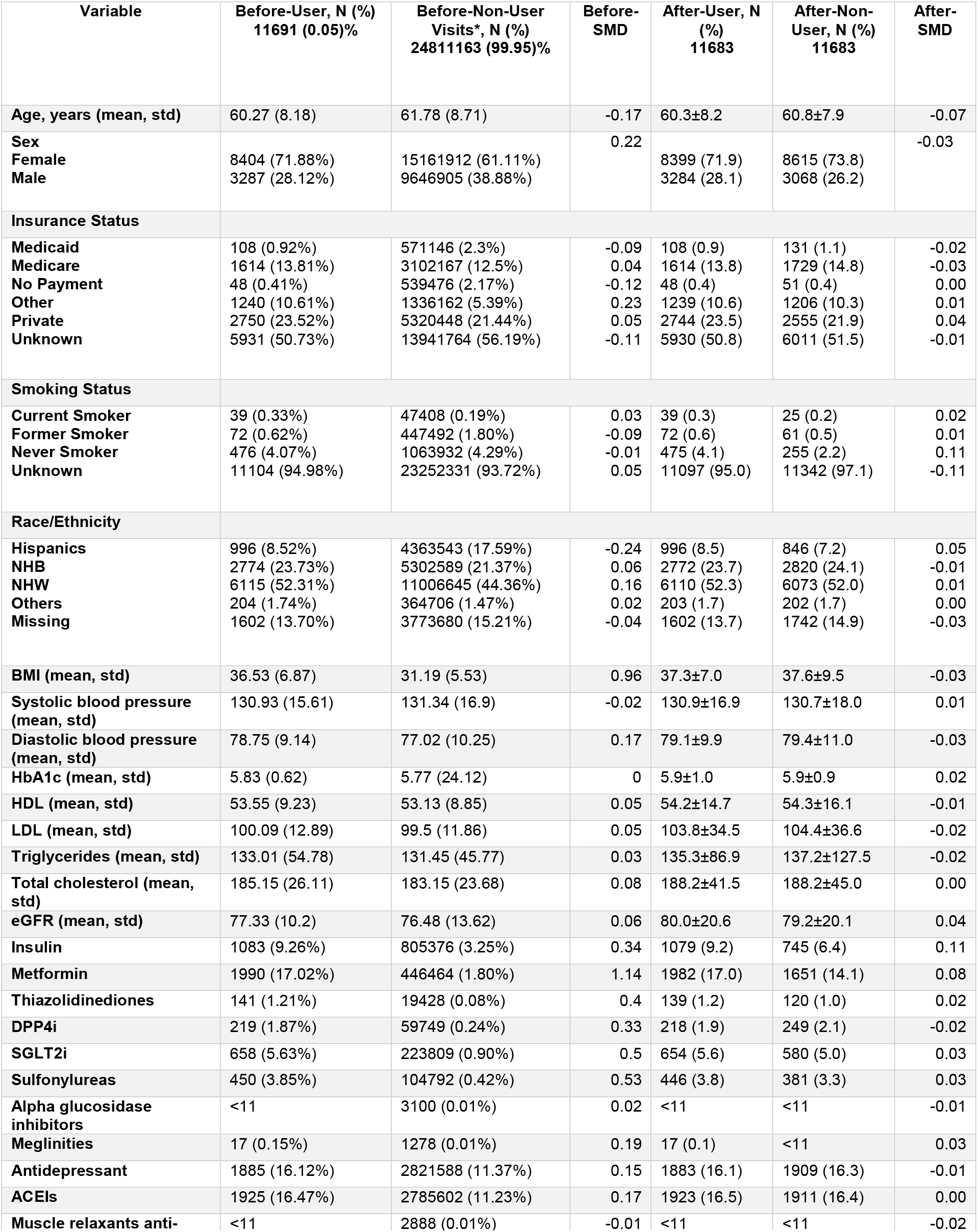
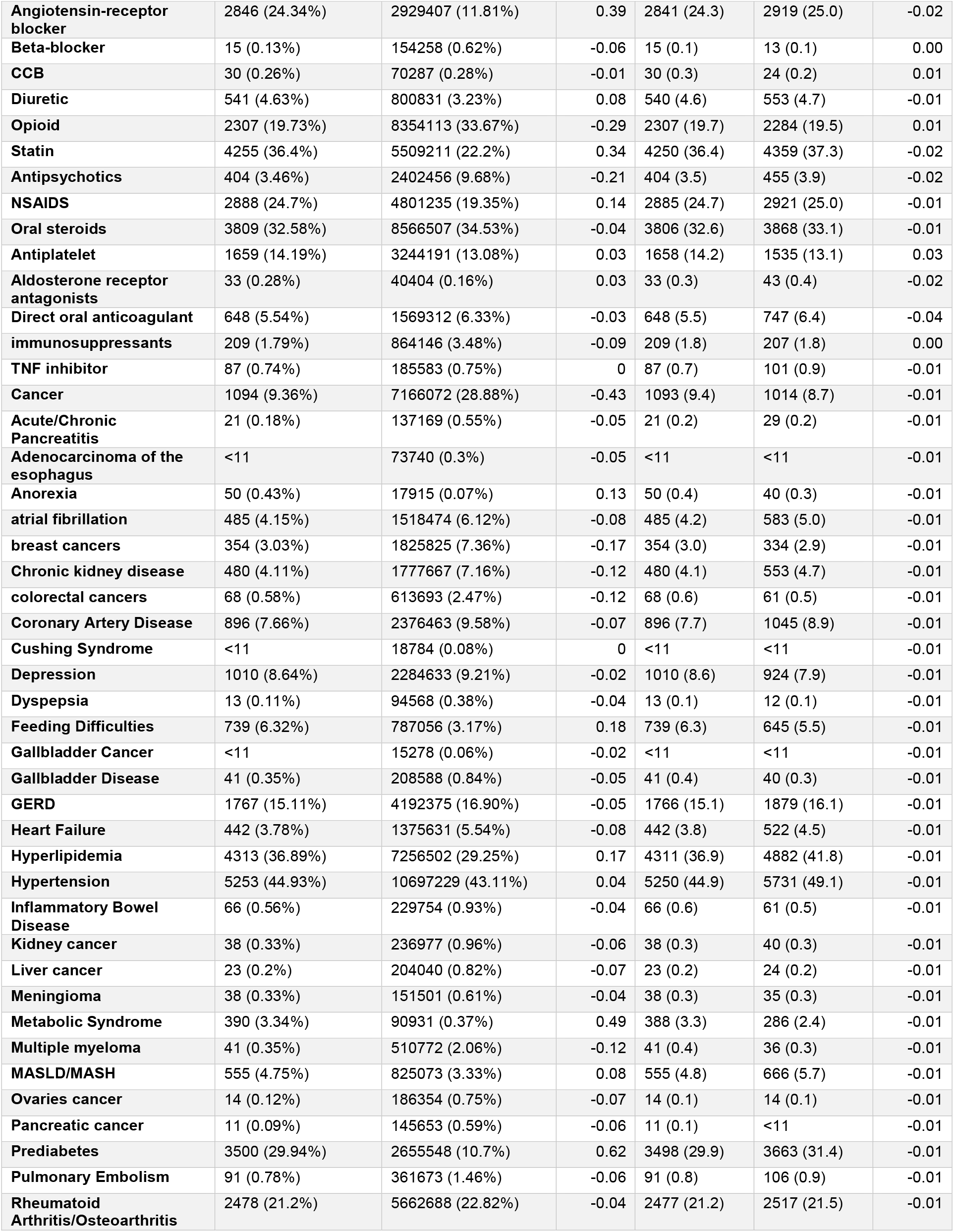
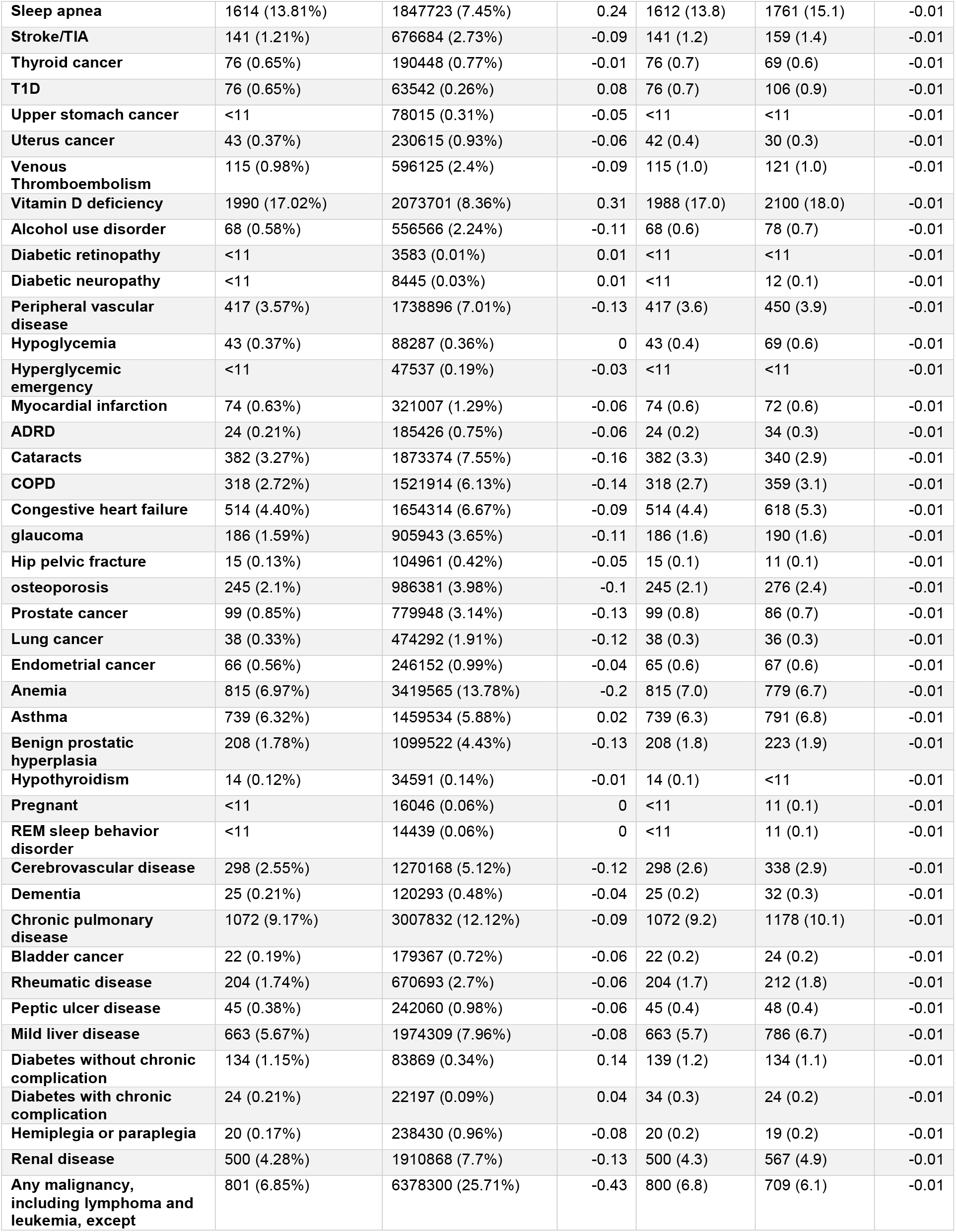
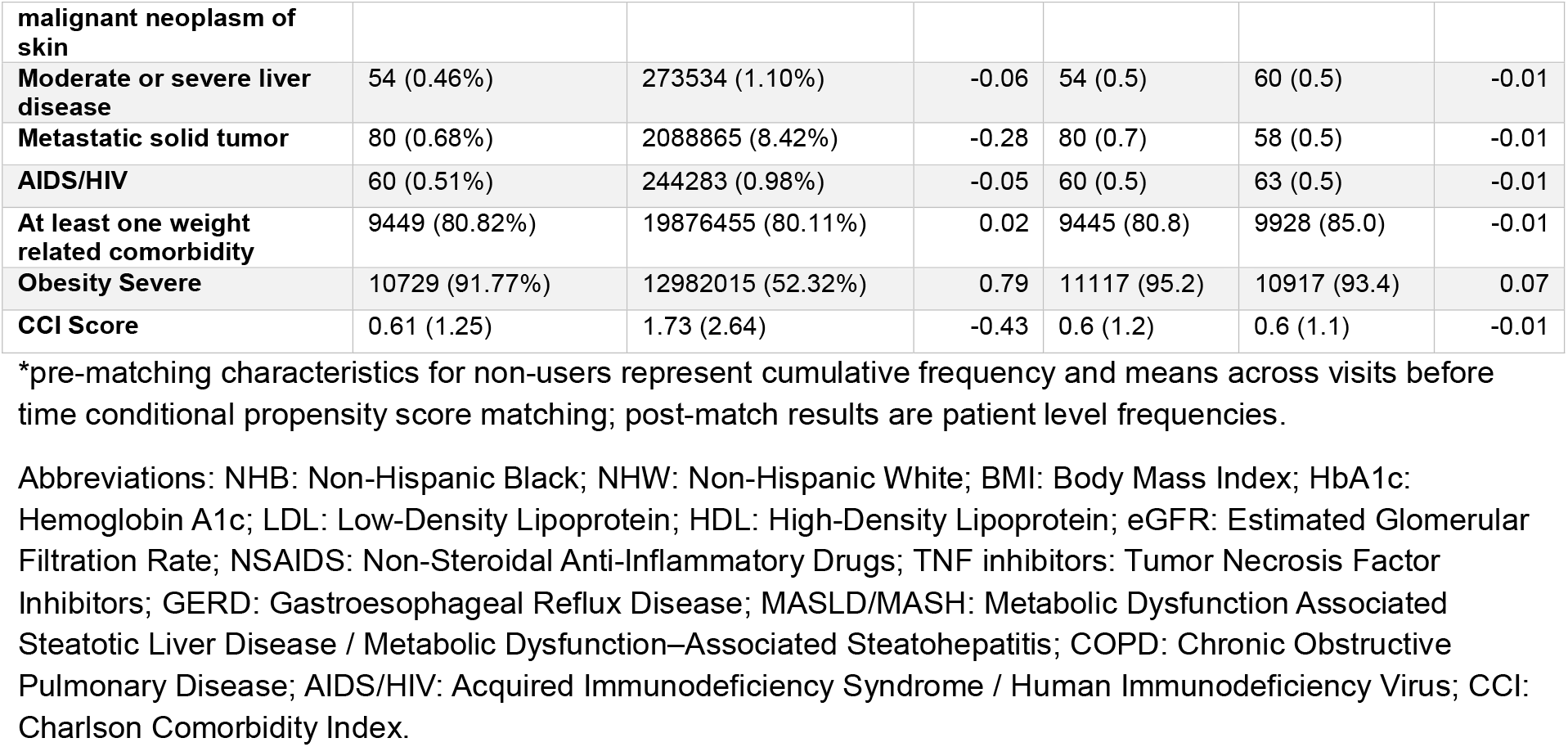
Baseline characteristics of GLP-1RA User vs. GLP-1RA Non-user cohort after 1:1 time conditional propensity score matching.

**Figure:**
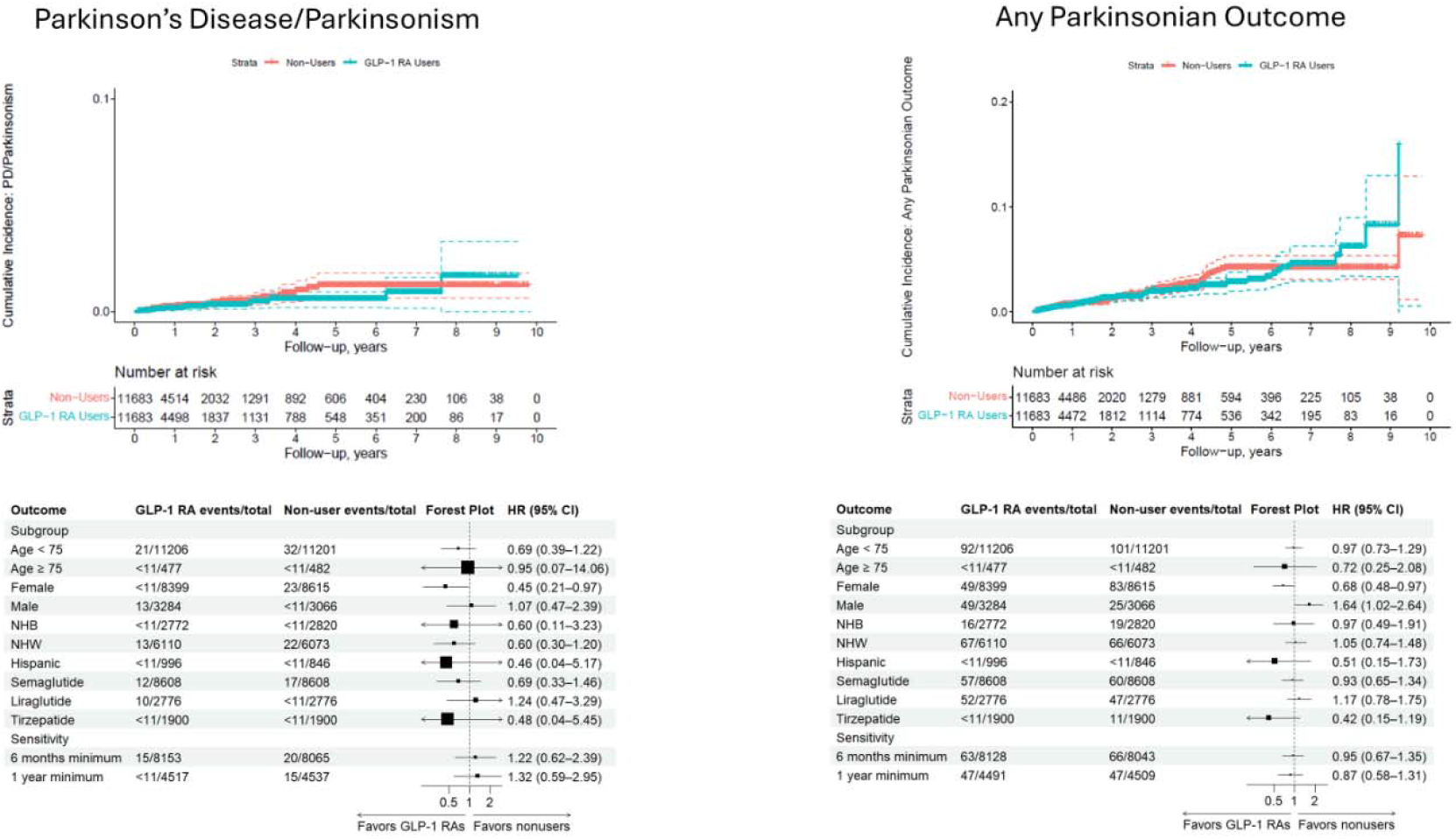
Cumulative Incidence and Subgroup Analyses of Parkinsonian Related Outcomes Among Patients with Obesity Using Glucagon-Like Peptide-1 Receptor Agonists (GLP-1RAs) vs Non-Users Abbreviations: NHB, non-Hispanic Black; NHW, non-Hispanic White.

## Data Availability

Data set Available through OneFlorida+ Clinical Research Network (email)

